# Ethnomedicinal health seeking practices and their associated treatment outcomes in managing diarrheal diseases among under-five-year-old children in Korogwe and Handeni Districts, Tanzania: Protocol for a cross-sectional mixed method study

**DOI:** 10.1101/2023.02.14.23285909

**Authors:** Edwin Liheluka, Sophia Nyasiro Gibore, John P. A. Lusingu, Samwel Gesase, Daniel T. R. Minja, Maike Lamshöft, Denise Dekker, Theodora Bali

**Affiliations:** Department of Public Health and Community Nursing, University of Dodoma, Dodoma, Tanzania; Department of Educational Psychology and Curriculum Studies, University of Dodoma, Dodoma, Tanzania; National Institute for Medical Research, NIMR, Tanga Centre, Tanga, Tanzania; Bernhard Nocht Institute for Tropical Medicine, BNITM, Hamburg, Germany; German Center for Infection Research, DZIF, Germany; University Hospital Hamburg-Eppendorf, Germany

**Keywords:** Ethnomedicinal remedies, diarrheal diseases, medicinal plants, under-five-year-old children

## Abstract

**Background:** Ethnomedicinal remedies relevant for treating a range of ailments including diarrheal diseases among children aged less than five years is an integral component of the long-standing culture that communities have inherited from previous generations. The treatment also has mutual impact on the practice of health seeking behavior built within the family and clan level.

**Study Objective:** To assess ethnomedicinal health seeking practices and their associated treatment outcomes for managing diarrheal diseases among children aged below five years children in Korogwe and Handeni Districts, Tanzania

**Methods:** A mixed method approach will be employed whereby both qualitative and quantitative research approaches will be utilized. Narrative and cross-sectional research designs will be used for qualitative and quantitative research, respectively. The study population will include caretakers of under-five-year-old children, pediatric health care workers and traditional healers. Purposive sampling method will be used to select participants for qualitative research while a systematic random sampling will be used to select participants for quantitative research. Social Ecological Model (SEM) theory will be employed to elicit the target population perceptions and context-specific factors, which will explain the ethnomedicinal health seeking practices and their associated treatment outcomes in managing diarrheal diseases among under-five-year-old children in Korogwe and Handeni Districts, Tanzania.

## Introduction

Globally, there has been a tremendous increase in the use of ethnomedicinal remedies in managing different ailments. It is estimated that, about 80% of the world population makes use of ethnomedicinal remedies in managing common illnesses, since modern medicines are perceived inaccessible, costly and unsuited [1]. For instance, in 2019 80-99% of the population in Benin and Congo-Brazzaville, utilized ethnomedicinal remedies [2]. The popularity of ethnomedicinal remedies has been reported even in the western world even though there is little historical tradition for those remedies and the main reasons cited for using them include the adverse outcomes of the modern medicines and the increase of chronic diseases such as cancer and cardiovascular diseases [2]. In Canada and Brazil for example, about 1-19% of the population utilizes ethnomedicinal remedies to treat different illnesses [2].

Various health-seeking behavior studies have shown that communities differ with regard to their preference of seeking diarrhea treatment [3, 4]. Some prefer to seek treatment from biomedical health workers, and others prefer to seek treatment from Traditional Healers (THs). Likewise, there are also those who prefer self-treatment and there are those who prefer to be treated by all the above-mentioned health practitioners [3, 4]. Literature revealed that ethnomedicinal remedies applicable for diarrhea treatment are reliably linked with people’s cultures and have been used in the past and are still used to date to treat diarrheal diseases in people of all ages, particularly in Low and Middle Income Countries (LMIC) [4-7].

According to WHO report, there were 1.7 billion cases of diarrhea in 2017, with 525, 000 of those cases resulting in death [8]. Significant numbers of diarrheal related deaths occur among children less than two years of age residing in limited resource settings [9]. It is argued that, the increasing utilization of ethnomedicinal remedies in treating childhood diarrhea is an integral part of the long-standing customs that communities have inherited from previous generations [6]. Studies have highlighted the therapeutic potential of ethnomedicinal remedies in treating mild and severe diarrhea for children aged less than five-year old [3, 5, 10].

In Tanzania diarrhea is one of the most important health problems among children, statistics indicate that, the overall prevalence of diarrhea in the country was 25 % in 2015, much higher (14%) in urban areas than in rural areas (11%) [11]. Diarrheal diseases are fundamentally preventable, treatable, and avoidable. The disease can be prevented by improving living conditions, such as access to safe drinking water, sanitation and hygiene [8]. However, the disease continues to claim lives of many people especially children aged below five years living in the poorest parts of the world [8]. There have been various efforts, which have been taken by the Tanzanian government to combat diarrheal diseases among children below five years of age. For example, the introduction of vitamin A supplement to children across the country in 1997 [12] and the introduction of rotavirus vaccine in 2013 [13] have been associated with a significant reduction in diarrhea-related illnesses in Tanzania. Studies have shown that ethnomedicinal remedies are also among the interventions, which are widely used to manage diarrheal diseases among under-five-year-old children. However, their contribution in curbing the ailment is not well known [5, 7, 14, 15].

In Tanzania, the use of ethnomedicinal remedies is rooted within the traditions and customs of many communities [4, 16]. It is projected that, about 60% of Tanzanian population self medicate by using ethnomedicinal remedies prior to seeking healthcare in the formal health care facilities [17]. In Tanga Region, most users of ethnomedicinal remedies do not disclose publicly whether they use the remedies to treat their children who are suffering from diarrheal diseases [18]. This leads to a lack of adequate information regarding the actual number of users of ethnomedicinal remedies [18]. However, a study conducted in Lushoto and Pangani Districts in Tanga Region revealed that about 70% of study participants prefer ethnomedicinal remedies over modern medicine in case they suffer from various diseases including diarrhea [4].

With regards to safety, ethnomedicinal remedies have been discovered and developed from different cultures and different settings which lead to the nonexistence of common methods for most of the remedies, which are well known to everyone on how to prepare and consistently deliver them in a standard and regulated manner [1]. Despite their popularity and promising potential in treating diarrheal diseases among under-fives children, ethnomedicinal remedies are still considered to have limited quality, formal guidelines and scientific merits compared to modern medicines [14, 19]. However, there are ongoing endeavors being pioneered by the WHO that aim to harmonize the production and delivery of ethnomedicinal products [2].

Studies have shown that, ethnomedicinal remedies used for diarrhea treatment have some adverse effects; including nausea, exhaustion, vomiting and stomachache [10]. Additionally, the remedies can cause “black tongue and tinnitus in humans and abortion and liver complications in animals” [20, 21]. In Tanga Region, even as the use of ethnomedicinal remedies is very common, there are limited studies that have assessed the treatment outcomes associated with the use of ethnomedicinal remedies. Therefore, in order to better inform policymakers and general public there is a need for generating data in this area in order to have a better assessment on how ethnomedicinal remedies would complement the formal health care delivery outlets in the fight against diarrheal diseases in the Region.

Therefore, the broad intent of the present study is to assess ethnomedicinal health seeking practices and their associated treatment outcomes for managing diarrheal diseases among under-five-year-old children in Korogwe and Handeni Districts, Tanzania. In order to accomplish the focal research goal, the following are the specific objectives:

1. To determine the proportion of ethnomedicinal remedies users for treatment of diarrheal diseases among under-five-year-old children in Korogwe and Handeni Districts, Tanzania.
2. To determine socio-economic factors associated with ethnomedicinal remedies use in treatment of diarrheal diseases among under-five-year-old children in Korogwe and Handeni Districts, Tanzania.
3. To determine socio-demographic factors associated with ethnomedicinal remedies use in treatment of diarrheal diseases among under-five-year-old children in Korogwe and Handeni Districts, Tanzania.
4. To assess the treatment outcomes resulting from the use of ethnomedicinal remedies in managing diarrheal diseases among under-five-year-old children in Korogwe and Handeni Districts, Tanzania.
5. To explore how ethnomedicinal diarrheal treatment is provided to under-five-year-old children in Korogwe and Handeni Districts, Tanzania.
6. To document ethnomedicinal plants used to treat diarrhea among under-five-year-old children in Korogwe and Handeni Districts, Tanzania.

The results of the study might provide vital information on the importance of ethnomedicinal remedies to the policymakers, which will lend a hand in formulating effective guiding principles that would possibly guide ethnomedicinal treatment practices in Tanzania. Furthermore, the findings may as well facilitate to recommend realistic social and cultural interventions in managing diarrheal diseases for under-five-year-old children that will abide to communities’ social and cultural norms, which are vastly respected.

### Conceptual Framework

This study will adapt the Social Ecological Model as its conceptual framework [22]. Existing literature shows that, SEM is relevant in identifying context-specific factors, which motivate caretakers to utilize ethnomedicinal remedies for diarrhea treatment for their under-five-year-old children [22]. The SEM is a multi-level model with overlapping layers of factors which influence human behavior.

The first level of the SEM is at the core and addresses individual factors. At this intrapersonal level, individual factors such as demographic characteristics (age and gender) as well as socioeconomic status may influence the decision-making on the use of ethnomedicinal remedies for managing diarrheal diseases among under-five-year-old children [23] as cited by Winch, (2012), [22].

The second level is the microsystem, which involves interactions between the parent/caretaker and other family members. At this interpersonal level, a caretaker’s decision on where to seek medical care is connected with social networks and social engagements [23] as cited by Winch, (2012), [22]. The SEM posits that family members or neighbors influence may encourage a parent/caretaker to utilize ethnomedicinal remedies to treat diarrheal diseases among under-five-year-old children [3]. Other micro system level factors include affordability of ethnomedicinal remedies, family and neighbor’s perceptions, beliefs and practices towards ethnomedicinal remedies for managing diarrheal diseases.

The third level is the mesosystem, which involves interaction between other distal factors beyond the family and/or neighbors. Examples might include religious leaders, traditional healers/or healthcare providers. The approval or disapproval of the use of ethnomedicinal remedies by faith leaders and acceptance by family members may influence their use or no use of ethnomedicinal remedies in treating their under-five-year-old children.

The SEM suggests that various beliefs related to a specific illness (e.g., diarrhea) in the respective communities may either positively or negatively influence the decision-making process in the selection of the type of treatment or medication in treating different illnesses [23] as cited by Winch, (2012), [22]. For example, if a particular form of diarrhea is presumed to be caused by superstition; some community members believe that this type of diarrhea cannot be treated with modern medicine and the only alternative is to use ethnomedicinal remedies [24].

The fourth level is the exosystem, which consists the organization of healthcare delivery services which influencing caretakers to seek help for treating diarrheal diseases among their under-five-year-old children [23] as cited by Winch, (2012), [22]. For example, distance of the health facility from where the caretakers live is likely to contribute to the use of ethnomedicinal remedies.

The fifth level is the macro system, which are the broader socio-cultural, policy and/or political environment [23] as cited by Winch, (2012), [22]. In the context of this study these include socio-cultural, policy and political environment such as recognition of traditional healers and their perceptions and practices towards treatment of diarrheal diseases for under-five-year-old children in Tanzania.

The SEM underscores that laws and policies in any locality can or cannot make an individual to be more flexible in choosing which medicines s/he prefers to use in treating diarrheal diseases for under-five-year-old children. For example, in Tanzania the health policy recognizes THs and their medicines, which are used to treat diarrheal diseases [17]. Additionally, literature suggests that there are some cultures that encourage the use of ethnomedicinal remedies in treating diarrheal diseases among under-five-year-old children [25].

## Methods

### Study setting

The present study will be carried out in Korogwe and Handeni Districts, which are among the eight Districts of Tanga Region, Tanzania. The two Districts have been earmarked for this research since they considerably embody the practices of caretakers utilizing ethnomedicinal remedies for the management of diarrheal diseases among under-five-year-old children[4, 26]. The remedies are also considered to be closely connected to people’s collective cultural life [4, 26].

According to the 2012 human population census survey, Korogwe District had an approximate population of 242,038 [27]. The main economic activities in the District include trade, fishing, farming and livestock keeping [28]. Handeni District has a population of 276,646 [27]. The vast majority of the populations in the District are farmers engaged in cultivation of maize, cassava and beans [28].

### Study Approach

A mixed method approach will be employed whereby both qualitative and quantitative research approaches will be utilized. A mixed method will enable the researcher to collect more comprehensive data and obtain in-depth understanding of the research problem than when using a standalone quantitative or qualitative research, as it incorporates advantages of both techniques [29].

### Qualitative research

A qualitative study approach will be employed to explore how ethnomedicinal diarrhea treatment is provided and to document ethnomedicinal plants which are used to treat diarrhea among under-five-year-old children. The approach is relevant because it has the potential to discover unknown or less reported research problems [30].

### Quantitative research

A quantitative study approach will also be employed to capture information on the proportion of ethnomedicinal remedy users for treatment of diarrheal diseases, socio-economic factors, socio-demographic factors and treatment outcomes related to the use of ethnomedicinal remedies in managing diarrheal diseases among under-five-year-old children. The approach is suitable for this study because data are collected methodically which allows the investigator to attain a bigger sample size which is vital in realization of candid comprehensive study conclusion [31].

### Study Designs

This study will employ exploratory sequential mixed method design whereby qualitative data will be collected and analyzed first. The quantitative analysis will be employed to determine relative prevalence of ethnomedicinal remedy use for managing diarrheal diseases among under-five-year-old children in Korogwe and Handeni Districts in Tanga Region.

### Qualitative research

The qualitative part of the present study will employ a narrative research design. The design has been favored because it permits the researcher to visualize matters that have never been reported elsewhere [32]. For example, the design will help the researcher to learn from study participants about their views on the use of ethnomedicinal remedies, where the remedies are found, how they are processed and prescribed.

### Quantitative research

The quantitative part of the present study will employ a hospital-based cross sectional study design. Quantitative research will only involve caretakers who will have brought their under-five-year-old children to be treated at the selected 12 health facilities earmarked for this study in Korogwe and Handeni Districts. For those caretakers whose children have had diarrhea in the past six months will have the opportunity to participate in the study. Cross-sectional study design is chosen since it allows the researcher to get a depiction of a definite faction of people at a particular point in time [33].

### Study Population

The study population includes caretakers of under-five-year-old children, pediatric health care workers and THs in Korogwe and Handeni Districts. The caretakers have been chosen since they are users of ethnomedicinal remedies, while health care workers and THs are responsible in managing diarrheal cases by using modern and traditional medicines, respectively.

### Inclusion criteria

Caretakers of under-five-year-old children living in Korogwe and Handeni Districts, attending health facilities (seven in Korogwe District and five in Handeni District). Traditional healers who treat under-five-year-old children in Korogwe and Handeni Districts and pediatric health workers (clinicians and nurses) working in the main 12 health facilities earmarked for this study.

### Exclusion criteria

Respondents who will be seriously sick and cannot participate in the study and respondents who will be absent on the day of data collection

### Data Collection Methods

#### Qualitative research

Focus Group Discussions (FGDs) and In-depth Interviews (IDIs) will be employed in this study to facilitate data collection. The methods have been chosen because they are the most common source of qualitative data in health research [34].

The guides will be used to interview THs, health care workers and caretakers of under-five-year-old children. The guide for these groups of participants will have two sections. The first section will collect information on general knowledge of diarrheal diseases and the use of ethnomedicinal remedies in managing diarrheal diseases among under-five-year-old children. The second section will collect information related to ethnomedicinal remedies that are used to treat diarrheal diseases among under-five-year-old children and the reasons why caretakers prefer to use those remedies. Participants will also be asked about the safety and efficacy of these remedies.

#### Quantitative research

A questionnaire will be administered to the study participants at the health facility during exit. The face-to-face interview will start once a study participant has consented to participate in the study and s/he has signed and dated the Informed Consent Form (ICF) (Appendix 6). The principal investigator (PI) or research assistant will explain to the study participant in details about the goal of the study. The questionnaire has three sections. Section one will collect information from caretakers that include age, sex, marital status, occupation, address, employment status, number of children in the family and education. The questionnaire will also collect household information (access to clean and safe water, main source of drinking water, location of the house, ownership and type of latrine and housing materials). Also, information on ownership of valuable properties (mobile phone, TV, radio, car, bank account, motorbikes, refrigerators, computer etc.) will be gathered. Section two will collect information from caretakers related to treatment outcomes after using ethnomedicinal remedies for example if the child has recovered or the child has experienced adverse effects after using ethnomedicinal remedies. Section three will collect information allied to health practises (health seeking behaviour, participation in health education and ownership of health insurance). The interview will take about 30 to 45 minutes to complete. Participant’s responses will be recorded manually in the questionnaire.

#### Sampling techniques of study participants and health facilities

Purposive sampling technique will be used to select FGD’s and IDI’s participants and 12 health facilities (seven in Korogwe District and five in Handeni District). Purposive technique has been chosen because it allows the researcher to select and include participants/institutions in a specific study who/which are knowledgeable and experienced about the study topic [34].

#### Selection of IDI participants

Once potential participants have been selected, the PI will look for participants in the areas where they live one week before the interview date and inform them that they have been selected to participate in the study. The PI will take time to enlighten each participant about the study objectives and the importance of their participation in the study. Similarly, the PI will obtain contact details from each participant including mobile phone number, house number, sub village name, etc, if at all participants will be having/knowing them in order to facilitate communication before and after the interview. Twenty-four hours prior to the interview date, the PI will reach all participants invited in that day in order to remind them again about the interview.

The study investigator with the help of a research assistant will facilitate all IDI’s. The interview process will start once respondent has agreed to participate in the study. Data will be recorded by taking notes; and where a participant agrees, a digital audio-visual recorder will be used. One in-depth interview will take approximately 30 to 45 minutes until completion. Only one researcher will be interviewing the IDI’s participant. A research assistant will be taking notes.

It has been suggested that while conducting in-depth interviews, saturation point occurs at least within the first 12 interviews [35]. Data saturation refers to the situation in the research procedures whereby there is no new information which is being reported by the respondent [35]. Taking into account of the aforesaid recommendation, the present study will recruit 24 health care workers, 24 caretakers whose under-five-year-old children have experienced diarrhea within the past six months and 44 THs as explained here under. However more IDIs might be added accordingly until data saturation point is realized.

Korogwe and Handeni Districts have a total 11 divisions whereby four divisions are in Korogwe District namely Mombo, Bungu, Magoma and Korogwe while Handeni District has seven divisions which are Chanika, Sindeni, Mkumburu, Magamba, Kwamsisi, Mzundu and Mazingara [36]. Within the two Districts there are only 12 main health facilities (four Hospitals and eight Health centers). The present study aims to recruit participants from each 11 divisions and each 12 main health facilities in the Districts. The aim is to gather a wide range of outlooks from stakeholders across the two Districts. Twelve main health facilities are serving most under-five-year-old children, diarrhea patients in the two Districts which is why they have been earmarked for this study [37].

#### Selection of paediatric health care workers

Two health care workers (one pediatric clinician and one pediatric nurse) will be purposively selected from each of the 12 main health care facilities to participate in this study, which will make the number to reach 24. Pediatric health care workers have been selected because they attend under-five-year-old children who are suffering from diarrhea and other diseases. Furthermore, they may be more aware of the different types of treatment that under-five-year-old children receive.

#### Selection of caretakers

Two caretakers (a female and a male caretaker) will be purposively selected from each of the main 12 health care facilities to participate in this study, which will make the number to reach 24. Caretakers who brought their under-five-year-old children to the health facility for diarrheal treatment for the past six months will be selected from the District Health Information System (DHIS) book to participate in this study. The investigator will locate the participants where they live and explain to them that they have been selected to participate in the study. The investigator will also explain in detail about the study and arrange the interview date with the participant. DHIS book includes a variety of patient information such as name, place of residence, gender, age and the disease that brought the patient to the health facility for treatment. Caretakers are included in this study because they are responsible for deciding what treatment is best for their children when suffering from diarrhea. Furthermore, this study proposes to have a female and a male caretaker in order to obtain diverse views from both sexes. Caretakers who brought their children to the hospital within the previous six months will be selected to take part in this study because they may still have a good memory of the diarrhea episode that occurred to their children.

#### Selection of THs

At least four THs will be purposively selected from each of the 11 divisions to participate in the present study. The TH coordinators of Korogwe and Handeni Districts will be consulted to select at least four THs from each division who will be included in the study, which will make the number to reach 44. The criteria for selecting THs will be their experience in treating children within Korogwe and Handeni Districts. THs have been included in the present study because they occasionally act as the first point of contact for under-five-year-old children with diarrhea or sometimes as the referral point once the child fails to recover after visiting the health care facility.

#### Selection of FGD’s Participants

Eligible participants for this study are caretakers who brought their under-five-year-old children to the health facility for diarrhea treatment within the past six months and were registered in the DHIS book.

Once potential FGD participants have been chosen, the PI will locate participants in the area where they live one week prior to the interview date and inform them that they have been selected to participate in the study. The PI will explain to each participant in details about the goal of the study and the importance of their participation in the study. The PI will also obtain contact details for each participant in order to facilitate the process of communication before and after the interview. Furthermore, twenty-four hours before the day of the interview the PI will contact again all participants invited in that particular day in order to remind them about the interview.

One FGD will last approximately between 30 to 45 minutes depending on the knowledge and interests of the participants on the study topic. The interview process will start once all FGD’s respondents have agreed to participate in the study. Data will be recorded by taking notes and where agreed, by participants, a digital audio recorder will be used. The PI will lead the discussion and the research assistant will take notes.

Venue for FGD’s meetings will be organized with assistance from the local village leaders. Transport costs for study participants to the meeting venue will be reimbursed by the present study. The FGDs will be conducted before the IDI’s because if there will be some contradictory matters arising from FGDs, there will be a chance to seek more clarification through IDI’s with key informants.

Literature suggests that, at least six FGD’s are sufficient to answer research objectives in most qualitative studies [38]. The present study will conduct a total 24 FGD’s, 12 groups will be for male caretakers and 12 groups for female caretakers.

#### FGD’s composition

According to Green and Nicki (2018) the ideal extent of a FGD is six to ten participants [34]. A FGD of more than ten participants is difficult to control. Therefore, based on the above acumen, this study will invite a minimum of six and a maximum of ten participants to each FGD.

#### Sample Size Estimate for Quantitative Component of the Study

A single population proportion sample size formula will be used to calculate the sample size [39]. A study conducted among caregivers in Dar-es salaam [10], reported 71% of participants used ethnomedicinal remedies for treatment of diarrheal disease for their under-five-year-old children; this study experience was taken to calculate the sample size. The desired level of Confidence Interval (CI) at 95%, a margin error of 5%, and non-response rate of 10% were included in the formula as follows: N = Z^2^ x P x (100-P)/E^2^ whereby N = Estimated Sample Size, Z = Standard Normal Deviation of 1.962 corresponding to 95% CI, P = Proportion of outcome under study, and E = Marginal Error at 5%.

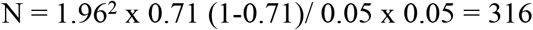

The final sample size is derived by the following formula: Final sample size = Effective sample size/ (1-non-response rate anticipated). The minimum sample size was calculated as 316, and the anticipated non-response rate or dropout was 10%. Then the minimum sample size = 316/ (1–0.1) = 351.9 ∼ (352)

#### Sampling procedures for quantitative research

The present study will employ a proportionate cluster sampling method. The sampling units are 12 main health facilities in Korogwe and Handeni Districts.

The estimated minimum sample size of 352 participants will be distributed based on the average of monthly diarrheal cases for under-five-year-old children reported in the 12 health facilities. To get the sample size for each health facility a total of average number of diarrheal cases for under-five-year-old children per month was calculated. To get the sample size for a respective health facility, the average number of the diarrheal cases per month was taken as a numerator and the denominator was the total number of all diarrheal cases of all health facilities and this was multiplied by the estimated sample size. For example, according to DHIS information the average number of diarrheal cases per month in February 2022 was 569 for both Districts (332 Handeni and 237 Korogwe). The average number of diarrheal cases per month for Magunga Hospital according to DHIS information was 76. Therefore, the estimated sample of caregiver to be selected from Magunga hospital was 76/569 × 352 = 47. This calculation was done for all 12 health facilities as presented in Table 1, which illustrates the number of participants that will be selected from each health facility based on the average number of diarrheal cases reported for under-five-year-old children per month.

**Table 1:**
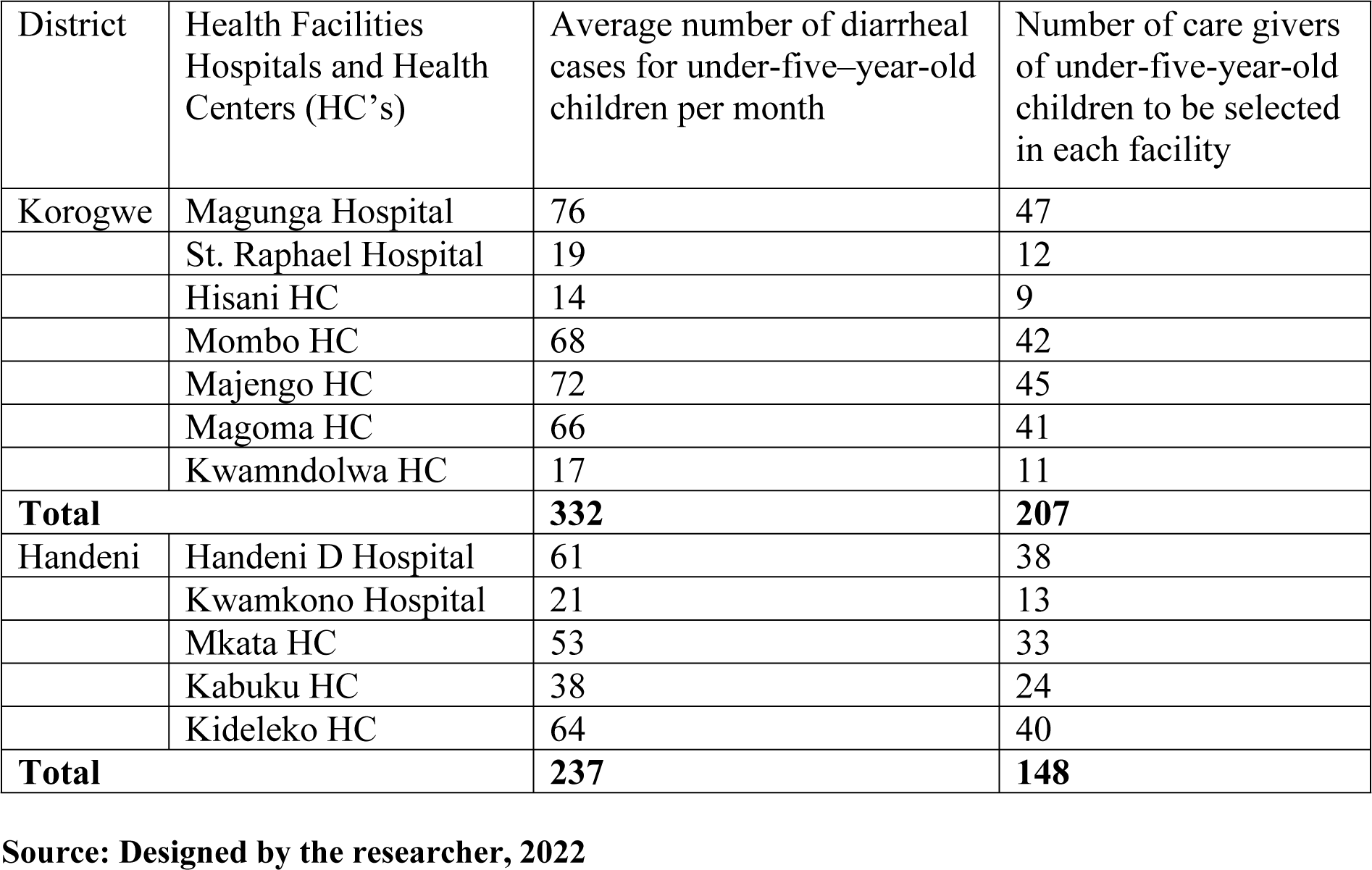
The average number of diarrheal cases per month for under-five–year-old children per month among 12 health facilities earmarked for this study in Korogwe and Handeni Districts

A systematic random sampling will be used to select eligible caretakers of under-five-year-old children attending the 12 health facilities as presented above. In order to systematically select eligible caregivers, a sampling interval will be calculated by using the following formula. The sampling interval (*n*th) will be calculated by dividing the estimated sample size (*n*) of the facility over the average number of diarrheal cases per month in the health facility (*N*). For example, the sampling interval (*n*th) for Magunga hospital will be 47/76 = 1. The first eligible caretaker will be selected blindly using a table of random numbers after which the remaining participants will be selected at regular intervals (*n*th). This process will be repeated until the required health facility sample size is achieved. The Principal investigator or research assistant will then contact the caretaker after his/her child has received treatment and invite him/her for the interview.

#### Data Collection Tools for Qualitative and Quantitative Parts of the Study

FGD and IDI guides with open-ended questions will be used to facilitate data collection for qualitative part of the research (Appendix 1-4), while a predesigned structured questionnaire with closed-ended questions will be used to collect quantitative data (Appendix 5).

### Study Variables and Measurements in the Quantitative study

#### Ethnomedicinal remedies

The primary dependent variable in this study is ethnomedicinal remedies use. The variable will be measured by asking caretakers if they ever used ethnomedicinal remedies for treatment of diarrheal diseases. The variable will be categorized into two groups as follows: 1= ever used ethnomedicinal remedies for treatment of diarrheal diseases, and 0= never used ethnomedicinal remedies for treatment of diarrheal diseases. This is the binary outcome, which assumes to follow binomial distribution i.e., 1 is the probability of success, in this case is ever used and 0 is the probability of failure, which is never used.

#### Treatment outcomes

Treatment outcomes is the secondary outcome which will be measured by asking caretakers about the therapeutic potential of ethnomedicinal remedies which might be positive outcomes like recovering from their illnesses or negative outcomes like not recovering from their illnesses or adverse events or death of the child after taking ethnomedicinal remedies. The variables will be categorized as 1= Recover from illness, 2= Not recover from illness, 3= Experiencing adverse effect after taking ethnomedicinal remedies and 4= others, specify

#### The independent variables comprise

I. Socio-Demographic Factors: these include all social and demographic characteristics of the respondents namely, age of the caretaker, sex of the caretaker, level of education, marital status, partner’s education level, occupation and number of living children.
II. Socio-Economic Factors: these include all factors that can affect and influence respondents’ financial status namely Socio-economic status (wealth index), which will be computed using principal component analysis (PCA) and categorized as 1=lowest, 2=low, 3=medium, 4=high and 5=highest. Other variables, which fall under this category include, affordability of ethnomedicinal remedies, which will be, categorized 1=Yes, 2=No; Access of ethnomedicinal remedies, which will be categorized as 1=Easy to access, 2=Not easy to access; Access to health facility (distance from heath facility); Attended health education (1=Yes, 2=No), ANC visits (1=Yes, 2=No), Number of ANC visits, and Health insurance ownership (1=Yes, 2= No).

### Data Management and Analysis

#### Qualitative data management and analysis

IDI and FGD interviews will be carried out in Kiswahili because it is the language spoken by most people in Korogwe and Handeni Districts. Collected data will be transcribed verbatim and then translated into English. The PI will thoroughly read the transcripts before handing them over to an independent and experienced qualitative researcher to review them again in order to confirm and approve the validity of the information. Finally, data will be analyzed manually using thematic analysis framework. Thematic analysis is a “qualitative methodical process for examining and reporting subject matters contained by the data set” [40].

Thematic analysis has been preferred in this study because of its supremacy that permits the PI to spawn the most recent notions that have been derived from the information collected [40].

Most approaches of thematic analysis entail several regular strides namely; getting used to the collected data, constructing preliminary codes, looking for themes, examining themes, describing themes and writing the final report [40]. The analysis steps are explained here under:

##### Orientation to the information collected

The PI will get accustomed to data gathered so as to appreciate the prosperity of the information with regard to the study objectives. Data will be repeatedly perused and scrutinized in order to grasp the scope of the arguments. At this step the PI will start identifying potential codes that will be used in the next step of analysis [40].

##### Making preliminary codes

This stage involves drafting of preliminary codes relevant to the study objectives and systematize them into a worthwhile category. The focus here will be on developing key points that will facilitate linking what was reported by participants in relation to the research objectives [40].

##### Finding potential themes

After data have been formally coded through the entire data set, the PI will organize diverse codes into prospective study themes and put together applicable hinted information within the branded study themes. The goal here is to produce ultimate themes relevant to the study objectives that will be used in the final report writing [40].

##### Examining themes

At this stage the themes will be reviewed and refined. Equally, the PI will check if identified themes have sufficient supporting data collected during the interview. At this stage, other themes can be modified or removed if they appear to have no supporting data but also other themes can as well be merged if they are representing something similar [40].

##### Describing themes

At this stage, themes will be scrutinized, assessed and improved further for the sake of making them ready for final data analysis. The PI will ensure that all identified themes have sufficient supporting data to defend the argument that will be presented. The themes will also be set to have a meaningful and coherent flow of elaboration relevant to the study objectives [40].

##### Writing final report

After all the themes have been comprehended and synthesized properly, the final step is to write a report or to scientifically tell the story of the information collected by following the sequence of the arranged themes. It is recommended to avoid repeating arguments that have already been written and the report should be able to present satisfactory evidence of the subject matter from the information collected [40].

#### Quantitative data management and analysis

Quantitative data will be cleaned and analyzed using Stata version 15 (StataCorp. 2017. *Stata Statistical Software: Release 15*. College Station, TX: StataCorp LLC). The distribution of the numeric variables will be checked using probably density plot and summarized using the measure of central tendency (Mean/Median) with the corresponding measures of dispersion (standard deviation/inter-quartile range). Categorical variables will be summarized using frequencies and percentages.

Frequency and percentages will be used to determine the proportion of ethnomedicinal remedies used for the treatment of diarrheal diseases among under-five-year-old children. Chi-square test will be used to compare the proportion of ethnomedicinal remedies’ use across explanatory variables, including socio-demographic characteristics, socio-economic status and administrative geographic locations. Frequencies and proportions will be used to identify treatment outcomes resulting from ethnomedicinal remedies use for the treatment of diarrheal diseases among under-five-year-old children.

A modified Poisson regression model with a robust standard error and log link function will be used to estimate the prevalence ratio (PR) and 95% confidence interval to determine social and economic factors associated with ethnomedicinal remedies use in the management of diarrheal diseases among under-five-year-old children. This model has no problem of non-convergence and overestimation of the risk ratio/prevalence ratio [41]. A multivariable Poisson regression model will be used to control confounders, whereby an adjusted prevalence ratio (APR) will be estimated. The Akaike Information Criteria (AIC) will be used to compare the regression model in which the model with the AIC will be considered a parsimonious model. All analyses will be based on a two-tailed significance level of alpha = 0.05.

#### Trustworthiness of study process and data

Trustworthiness of a study “refers to the scale of assurance in information and methods used to ensure the quality of a research” [42] as cited by Connelly et al., 2016 [43]. There are four trustworthiness principles namely, credibility, dependability, confirmability and transferability [42] as cited by Connelly et al., 2016 [43].

Credibility: “refers to the assurance in the genuineness of the study results obtained” [45]. In order to ensure the legitimacy of the results obtained, the present study will execute the scientific research methods, which have been proposed earlier in the methodology section. Execution of scientific research methods is key in justifying the authenticity of the study results obtained [46]. Study participants who have liberally accepted to participate in the study have a better chance of giving worthy and credible information, something which is fundamental in substantiating the genuineness of the results obtained [46].

Furthermore, in order to establish the credibility of the study findings, the member checks concept will be applied in this study. After the data have been collected, transcribed and translated, a few participants will be consulted and given transcripts that apply to them to read. The goal is to confirm from the participant if what is written is basically what s/he said during the interview. By being able to consider the member checks concept, the findings of this study may be credible as recommended by credibility decree [46].

Transferability “refers to the circumstances of showing that study results obtained from one setting can also be obtained in a different setting if conducted by a different researcher” [45]. In order to ensure transferability, the present study has provided detailed information about sample size calculation and data collection methods that will be employed. Additionally, the present study clearly states the selection criteria of participants as well as the study settings. Adherence to this principle is also key in facilitating the process of determining the legality of the study findings should other research of this kind be conducted elsewhere by a different researcher [46].

Dependability: “refers to the circumstances of demonstrating that, the study results are dependable and could be repeated if conducted in a similar circumstance” [45]. For this to be achieved the present study will sufficiently describe the entire flow of the research conduct from the beginning to the very end until the results are obtained. Wholly describing the study conduct is essential in providing another researcher the framework for carrying out similar research and be able to obtain consistent results [46].

Confirmability: “refers to the extent of objectivity to which study conclusion is produced by what participants have reported and not the investigator’s curiosity” [45]. In order to ensure that the findings of this study are based on what participants have reported, the PI will prepare an “audit trail” which will demonstrate step by step how the information gathered will be able to bring about the results obtained. The sequence of steps used to achieve the study results assists the assessors or readers to substantiate the authenticity of the data collected as directly relating to the results obtained [46].

#### Ethical clearance and consent to participate

The present study was submitted to the UDOM Institutional Research Review Committee (UDOM IRRC) and received ethical approval (Reference number MA.84/261/02/ dated 24^th^ May 2022). The study also received approval from the office of the Tanga Regional Administrative Secretary (RAS) (Reference number RM/R.20/I VOL.III/61 dated 07^th^ June 2022). Study objectives and other relevant study information such as study setting, procedures, benefits, risks, duration, approval documents, confidentiality of the information collected and the like, were fully explained and presented to the Regional, District as well as Village authorities.

Potential participants will be selected after obtaining ethical clearance and permission from all relevant authorities. Before enrolment, eligible participants will also be given detailed information about the study as explained above. Potential participants will then be given a chance to ask questions and the PI will answer the questions accordingly.

Furthermore, participants will be informed that, their participation in the study is voluntary and they are free to decide whether or not to participate. Participants who will agree to participate in the study will be required to sign and date the ICF before study procedures begin.

## Discussion

Products derived from specific medicinal plants such as leaves, barks, roots, seeds and fruits are said to be a great source of medicines for both humans and animals and have been used for years to solve various health ailments [47]. It is estimated that about 40% of pharmacological factories rely solitarily on medicinal plants for essential raw materials that are used to produce medicines [48]. Due to the sensitivity and the splendid value of ethnomedicinal remedies which are resulting from medicinal plants, this study aims to determine ethnomedicinal health seeking practices and their associated treatment outcomes for managing diarrheal diseases among under-five-year-old children in Korogwe and Handeni Districts, Tanzania.

As mentioned earlier on, ethnomedicinal remedies have become popular and seem to be widely trusted and used by the majority of people worldwide [1]. In Tanzania, the use of the same remedies dates back to pre-historic and pre-colonial times since the existence of the first human where the remedies were the only treatment option available to serve people [49].

In Tanga Region where the present study will be carried out, the use of ethnomedicinal remedies in treating various ailments is exceedingly a popular practice [4]. One of the factors cited as contributing to the widespread use of ethnomedicinal remedies is the rich wealth of medicinal plants, which are found within the Region [26]. The eastern curve of Amani and Muheza highlands is one of the only 20 natural geographical settings in the world, which is branded by miscellaneous range of medicinal plant species [26]. In addition, the presence of a large number of THs whose ratio to population ranges between 1:343 in urban areas and 1:146 in rural areas has promoted the use of ethnomedicinal remedies in the Region [26]. To our knowledge despite the wide spread use of ethnomedicinal remedies in Tanga Region, no study has been conducted in the Region to assess the proportion of ethnomedicinal remedies use for treatment of diarrheal diseases.

This study wishes to determine the proportion of ethnomedicinal remedy users. By so doing, the study results might provide information to the policymakers, which will aid in formulating effective guiding principles that would possibly guide ethnomedicinal diarrheal treatment practices within Tanga Region and Tanzania in general.

On the other hand, although, ethnomedicinal remedies have been frequently used for managing diarrheal diseases among under-five-year-old children in Tanga Region, to our knowledge the outcome of this intervention has never been documented and diarrhea incidences still remain high. For example, diarrheal prevalence among under-five-year-old children in Tanga Region was 53% in 2017, 56% in 2018, 60% in 2019 and 59% in 2020 [44]. Therefore, the present study is envisaged to document the treatment outcomes following treatment of diarrheal cases with ethnomedicinal remedies among under five children. Getting in-depth information on the treatment outcomes might help to guide the implementation of cordial interventions that aims to combat diarrheal diseases within the Region and Tanzania in general.

Reviewed literature identified a few medicinal plants which are thought to be relevant in treating diarrheal diseases among under-five-year-old children in Tanga Region [4]. However, to our knowledge, the process of obtaining plant raw materials that are used to treat diarrheal diseases, the procedures of preparing them, the dosage and storage of those ethnomedicinal remedies/concoctions have not been well documented. The present study apart from addressing the above knowledge gaps, will also investigate other medicinal plants, which are used to treat diarrheal diseases. Missing this information would perhaps deny policy makers and the general public an opportunity to receive research feedback, which could potentially assist in formulating successful interventions in controlling diarrhea-related illnesses in Tanzania. If proven useful, the knowledge will complement the existing formal interventions from the modern medicine in curbing diarrheal related morbidities and mortality in the area.

### Study limitations

Interviews for this study will be conducted in Kiswahili because it is a language spoken and understood by a large number of people in the study area. During translation process of transcripts from Kiswahili to English some of the words might be skipped or missed. The PI will hire and work together with two experienced qualitative researchers who are fluent in both languages – i.e., Kiswahili and English. Therefore, this challenge will no longer be an obstacle.

On the other hand, a bias may emanate in the PI’s purposive selection of information-rich study participants. To avoid this challenge, the PI will involve District THs coordinators during the exercise of selecting THs. Moreover, research assistants will be involved during the exercise of selecting health care workers and caretakers, thus minimizing a possibility of this challenge being a hindrance.

## Data Availability

Research data will be made publicly available when the study is completed and published.

## Supporting Documents

Appendix 1-4: In-depth Interview and Focus Group Discussion guides

Appendix 5: Questionnaire

Appendix 6: Informed Consent Form

Appendix 6: Study Timeline

## Authors’ Contributions

EL, TB, SNG, conceptualized the study. EL, TB, SNG, JPAL, SG, DTRM are currently organizing field activities and data collection. EL, TB, SNG are conducting the study and will analyze the data while EL, TB, SNG, JPAL, SG, DTRM, ML and DK drafted, reviewed, edited the manuscript. All authors read and approved the final version of the manuscript.

## Acknowledgments

The authors would like to express their heartiest gratitude to the German Research Foundation through collaboration between NIMR and Bernhard Nocht Institute for Tropical Medicine, BNITM, Hamburg, Germany for funding the study. The authors also thank the authorities of Tanga Region (The office of Regional Administrative Secretary) and Districts of Korogwe and Handeni for allowing and helping the research work to continue without interruption. The authors would like also to thank all participants for taking part in this study. University of Dodoma is thanked for granting ethical approval to conduct the study and leading all technical and logistical matters related to this study. Appreciations also go to NIMR management for continuing to provide administrative support during the implementation of this research. Finally, the authors are grateful to NIMR Korogwe Research Station staff for providing assistance during data collection.

